# Leveraging artificial Intelligence and online psychotherapy to achieve efficient and coordinated services within a healthcare setting: A quality improvement initiative

**DOI:** 10.1101/2025.02.22.25321970

**Authors:** Callum Stephenson, Jazmin Eadie, Christina Holmes, Kimia Asadpour, Gilmar Gutierrez, Anchan Kumar, Jasleen Jagayat, Charmy Patel, Saad Sajid, Oleksandr Knyahnytskyi, Megan Yang, Taras Reshetukha, Christina Moi, Tricia Barrett, Amirhossein Shirazi, Vedat Verter, Claudio Soares, Mohsen Omrani, Nazanin Alavi

**Author notes:** **Corresponding Author:** Nazanin Alavi MD FRCPC, Department of Psychiatry, Queen’s University; Hotel Dieu Hospital, 166 Brock Street, Kingston, Ontario, Canada K7L 5G2, Contact, 1-613-544-3310.

## Abstract

**Objectives:** This study aimed to implement an artificial intelligence-assisted psychiatric triage program, assessing its impact on efficiency and resource optimization.

**Methods:** This quality improvement initiative recruited patients on the waitlist for psychiatric evaluation at an outpatient hospital. Participants (n=101) completed a digital triage module that used natural language processing and machine learning to recommend a care intensity level and a disorder-specific digital psychotherapy program. A psychiatrist also assessed the same information and the decisions for care intensity and psychotherapy programs were compared with the artificial intelligence recommendations.

**Results:** The overall wait time to receive care decreased by 71.43% due to this initiative. Additionally, participants received psychological care within three weeks after completing the triage module. In 71.29% of the cases, the artificial intelligence-assisted triage program and the psychiatrist suggested the same treatment intensity and psychotherapy program. Additionally, 63.29% of participants allocated to lower-intensity treatment plans by the AI-assisted triage program did not require psychiatric consultation later.

**Conclusions:** Using artificial intelligence to expedite psychiatric triaging is a promising solution to address long wait times for mental health care. With future accuracy refinements, this could be a valuable tool to implement in hospital settings to assist care teams and improve mental health care. This could result in increased care capacity and improved workflow and decision-making.

## Introduction

Approximately 6.7 million Canadians experience mental health challenges annually, with half reporting unmet or partially unmet needs^1–6^. This contributes to a financial burden of over fifty billion per year^4,5^. The COVID-19 pandemic has exacerbated the strain on mental healthcare systems, leading to longer wait times, increased emergency department (ED) visits, and higher costs^1^. On average, patients in Canada wait nearly seven months to begin psychiatric treatment^7^. In the absence of timely outpatient care, many turn to EDs, which resulted in a 50% rise in mental health-related ED visits, in Ontario, from 2011 to 2021^8,9^. Nearly half of these visits are first contact, highlighting barriers to appropriate outpatient services^10–12^. ED care is also costly, at $500–600 a visit compared to $80–150 per visit in outpatient settings^8,13^. Furthermore, untreated mental health issues increase hospital stays, readmissions, and chronic disease management costs by 1.5 to 3 times^14–20^.

The Kingston-Frontenac-Lennox and Addington regions face similar challenges. Kingston Health Sciences Center (KHSC), which serves 650,000 individuals annually, had 1,200 individuals on its outpatient psychiatry waitlist and had wait times of 12 to 16 months in October 2023 to see a psychiatrist. In 2022, 9.02% of KHSC ED visits (4,760) were mental health-related, with 73.53% of these cases being redirected to outpatient care. Improving access to outpatient services could have prevented most of these visits, saving up to $1.4M/year and reducing overall ED demand by up to 6.63%.

To optimize resource allocation, Ontario employs stratified and stepped-care models, prioritizing psychiatrist appointments for severe cases^21–24^. However, current triage processes, such as those at the Mental Health and Addiction Clinic at Hotel Dieu Hospital (HDH), require a 45-minute interview conducted by a registered nurse before they can deliberate with the healthcare team and assign care. Although effective, these approaches are resource-intensive and prolong wait times^21–24^.

To address these inefficiencies, the research team developed an artificial intelligence (AI)--assisted triage program that uses natural language processing (NLP) to analyze patient narratives, referrals, and questionnaires (International Patent System (IPS); PCT/US22/43514). The AI-assisted triage program recommends care levels and suggests disorder-specific digital psychotherapy programs, enabling data-driven prioritization^25^. A transformer-based classifier trained on over 5,000 narratives from public forums (e.g., Reddit) achieved 90% accuracy in predicting symptomatic phrases compared to expert human agents^26,27^. Additionally, these algorithms have analyzed over 15 clinical trials with approximately 2,000 patients to predict outcomes for digital psychotherapy (i.e., assignments, feedback, symptom severity scores, engagement) and flag critical issues such as suicidal ideation^24,25,28–41^. By identifying key variables such as text composition and symptom severity, the AI-assisted triage program has been able to predict patient dropout rates with 70% accuracy, four weeks in advance, and can tailor treatment recommendations accordingly^26^. Using psychotherapy completion probability and symptom severity scores, through psychometrics like the Patient Health Questionnaire, a decision-making algorithm was developed to improve outcomes^42^. This results in low-intensity care are suggested for patients with moderate scores (PHQ-9<19) and high completion probability^26^. In contrast, high-intensity interventions were recommended for individuals with severe symptoms and/or low engagement^26^.

This quality improvement project was implemented to optimize outcomes, expedite care allocation, and reduce wait times^43–46^. Using this AI-assisted triaging could allow for the development of specialized treatment pathways and enhanced care delivery.

## Materials & Methods

### Participants & Recruitment

Participants on the outpatient waitlist for the Mental Health and Addiction Care, General Stream at KHSC (HDH site), referred between January 1 and September 30, 2023, were contacted. The program ran from October 2023 to March 2024 as part of a six-month funding period. This timeline was selected to avoid disrupting regular clinic triage and to focus on recent referrals for accurate data.

### Inclusion & Exclusion Criteria

Inclusion criteria included adult patients on the outpatient waitlist for KHSC’s Mental Health and Addiction Care, General Stream (January 1–September 30, 2023), residing in Ontario, proficient in English, and with reliable internet access to complete the AI-assisted triage and digital psychotherapy programs. Participants were informed this was not a crisis resource, and therapists were not immediately available. Those in a mental health emergency were not enrolled but directed to emergency resources (e.g., ED access, crisis lines), with incidents reported to the principal investigator (PI), and a psychiatrist. Participants’ health and comfort were prioritized, and they could withdraw anytime while remaining on the waitlist. If a crisis arose during the program, they would be promptly seen by a psychiatrist.

### Ethics Approval

This project received approval from the Queen’s University Health Sciences and Affiliated Teaching Hospitals Research Ethics Board (File: 6041568).

### Procedure

Participants were phoned by a research coordinator to assess their interest in participating. If interested then verbal consent was obtained and participants were sent a link to create an account on the secure cloud-based digital mental health platform, Online Psychotherapy Tool (OPTT) ^47,48^. Clients would then complete the AI-assisted triage module on OPTT which included psychoeducation material explaining the journey of someone with mental health problems and the struggles they might experience. Patients were then prompted to share their stories, concerns, and symptoms in detail. Participants could use typed or voice-inputted answers to share their stories. The triage module also included various questionnaires to assess symptom severity including the PHQ-9, Generalized Anxiety Disorder - 7 Item (GAD-7)^49^, and Ask Suicide Questionnaire (ASQ) to assess suicide risk^42, 49–50^. High-risk participants were identified, and a psychiatrist and nurse were notified and contacted the participant. Additionally, OPTT provided high-risk clients with crisis resources and directed them to the ED. The AI-assisted triage program analyzed the module and produced a report for the clinical team including the participant’s answers, questionnaire scores, and recommended care intensity and psychotherapy module (Figure 1).

**Figure 1.**
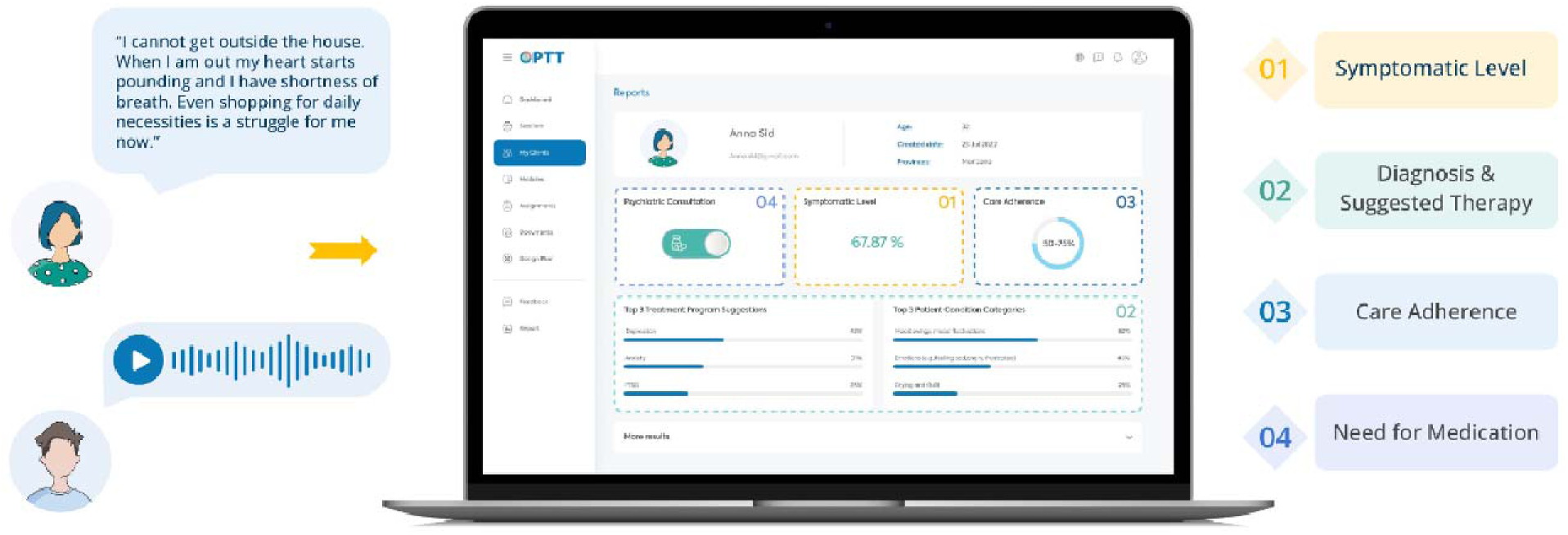
In the triage module, the patient is prompted to explain their mental health challenges using text or voice. Based on their answers, OPTT’s analytic algorithm assesses the most sensitive and symptom-related content, facilitating the review process for clinicians. Based on the nature of the problems explained in the patient’s narrative, the algorithm provides (1) a report about how symptomatic the patient is, (2) suggests an appropriate disorder-specific digital psychotherapy program, (3) provides a prediction on the patient’s adherence to care (i.e., care completion probability), and (4) the suggested care intensity level (4).

To evaluate the algorithm’s decision accuracy, a psychiatrist was used as the human comparator, as there are no established, validated, and evidence-based triage systems to evaluate the decisions. Therefore, all information collected from the triage module was reviewed independently by a psychiatrist to make recommendations for care intensity and psychotherapy programs, based on the Diagnostic and Statistical Manual of Mental Illnesses, 5th Edition (DSM-5)^51^. Once the participants’ treatment intensity and psychotherapy modules were confirmed, they were paired with their care team (Figure 2).

**Figure 2.**
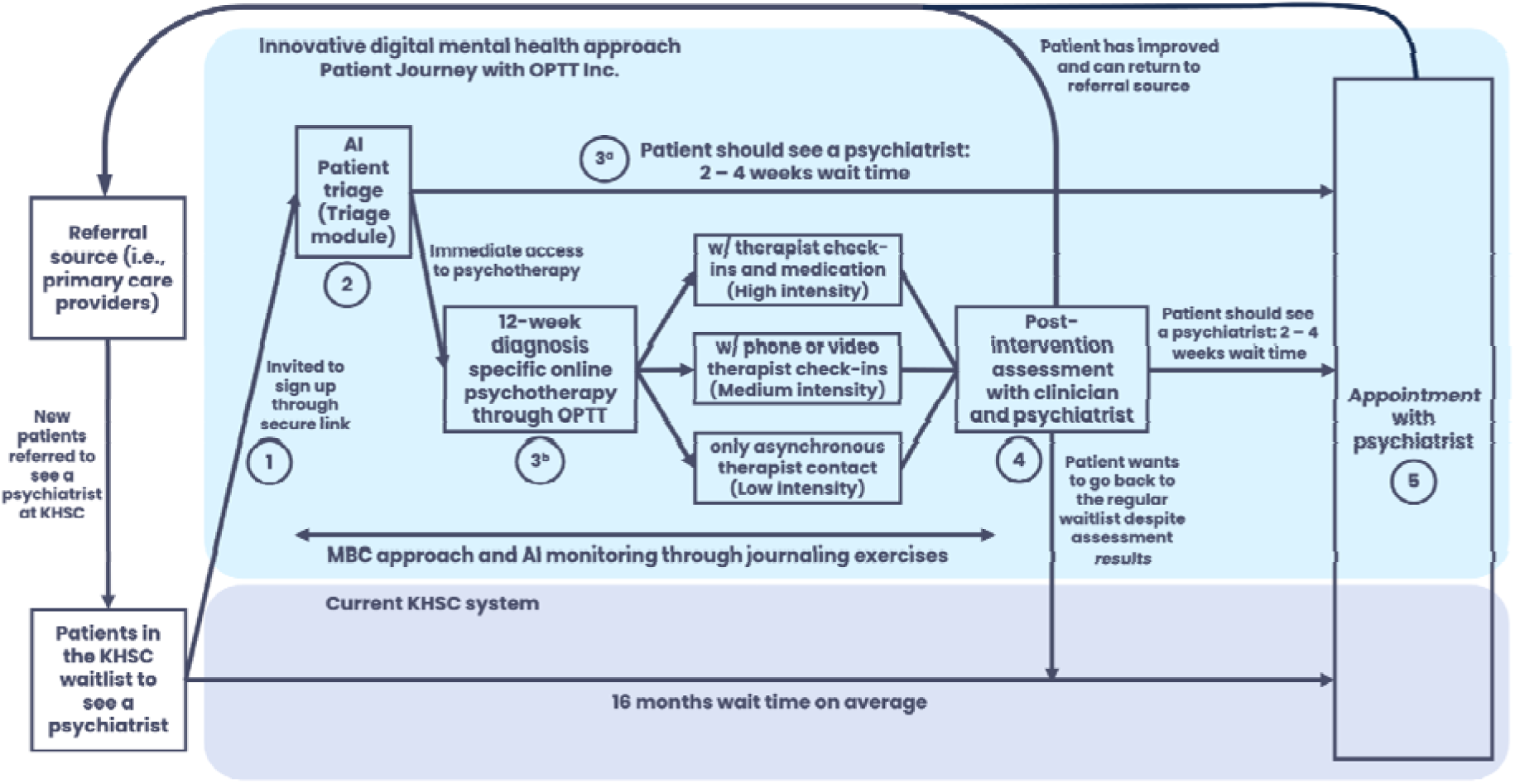
The participant journey from initial referral to psychiatric appointment.

### Platform & Technology

The platform (OPTT) used in this study provides a comprehensive range of digital care solutions, including an AI-assisted triage program which is powered by a proprietary NLP algorithm. Additionally, OPTT offers various online psychotherapy programs for different psychiatric conditions that facilitate data-driven decision-making and streamline care delivery. In this study, the algorithm categorized participants based on symptom severity into four groups:

1. *Mild (Level 1):* Scheduled to see a psychiatrist within six months.
2. *Moderate (Level 2)*: Scheduled to see a psychiatrist within three months.
3. *Severe (Level 3)*: Scheduled to see a psychiatrist within one month.
4. *Inconclusive (Level 4)*: The AI-assisted triage program or the psychiatrist could not allocate due to a lack of information from the participant and/or referral notes. Participants were scheduled to see a psychiatrist within one month and then allocated.

All participants were offered clinician-supervised, disorder-specific, online psychotherapy programs (∼10-12 weeks) while they awaited their psychiatrist appointments^24,25,28–41^. This ensured participants received timely care and therapeutic intervention, addressing immediate needs and optimizing care delivery. The type of treatment offered was chosen by the AI-assisted triage program based on symptoms (i.e., generalized anxiety disorder (GAD), major depressive disorder (MDD), dialectical behavioural therapy (DBT), posttraumatic stress disorder (PTSD), insomnia, substance use disorder (SUD), obsessive-compulsive disorder (OCD), COVID-19). The level of clinician involvement in the treatment plan was decided based on their assigned severity level:

1. *Mild (Level 1):* Therapist-guided, text-based online psychotherapy.
2. *Moderate & Severe (Levels 2 & 3)*: Therapist-guided, text-based online psychotherapy and a weekly phone/video call for additional support.

In some instances, the severity level was categorized as Level 1, but no suitable psychotherapy modality was available to address the participant’s specific needs (e.g., attention deficit hyperactivity disorder (ADHD)). Consequently, while their severity level was determined to be Level 1, the psychotherapy module recommendation was inconclusive. These patients were booked for an assessment with the psychiatrist to choose the most suitable course of action.

At the end of treatment, the therapist would discuss the participant’s care and progress with the psychiatrist. If it was determined that the patient no longer needed to see the psychiatrist, this would be communicated to the patient and their referring clinician, and the patient would be transferred back to their referral source for ongoing care (Figure 2). If the psychiatric intervention was still deemed necessary at the end of the psychotherapy program, the patient would be booked for their psychiatric appointment as planned.

### Online Psychotherapy

All psychotherapy modules mirrored their in-person counterparts, developed by the PI, an expert in the delivery of digital psychotherapy^52^. The previously validated modules included 20–30 weekly slides of psychotherapeutic concepts and strategies delivered asynchronously^24,25,28–41^. Participants received one session weekly via OPTT, with instructions to review materials and complete homework exercises to practice techniques and reflect on thoughts and symptoms^47,48^.

Each participant had a designated care provider trained in the delivery of digital psychotherapy, who reviewed homework and provided personalized text-based feedback. Feedback included validation, progress assessment, strategy suggestions, and key concept summaries in a signed digital letter. Session-specific feedback templates were used to ensure consistent, efficient feedback while maintaining personalization for each client. Participants and therapists communicated asynchronously on OPTT for non-therapy-related questions, with therapists responding weekly. For technical issues, OPTT’s support team was available^47,48^.

Participants in Levels 2 and 3 had weekly check-in calls with their therapist, through phone or video calls, depending on the client’s preference. Participants in Level 3 were seen by a psychiatrist who could prescribe medication following standard clinical guidelines.

### Outcomes

The following variables were used to evaluate the efficacy of the program:

1. *Wait times:* Changes in wait times to receive care and therapist time commitment as recorded by the research team.
2. *AI-assisted triage program decision-making accuracy:* Comparing the decision-making of the AI-assisted triage program (treatment intensity and psychotherapy program) to a psychiatrist.
3. *Care sufficiency:* Number of participants in Levels 1 and 2 who no longer required a psychiatric appointment after completing the psychotherapy program.

### Statistical Analysis

All analyses were conducted using IBM SPSS Statistics for Mac, version 25 (IBM Corporation, Armonk, NY, USA) with a two-tailed significance level of p<0.05. Decision-making accuracy between AI-assisted triage programs and psychiatrists was evaluated using a contingency table comparing classifications (mild, moderate, severe, inconclusive) or suggested treatments. The proportion of agreement was calculated as the sum of diagonal matches divided by total cases. Chi-square tests assessed whether AI classifications differed significantly from expected distributions under the assumption of no relationship with human classifications. A significant p-value indicated a statistically significant association between human and AI classifications. Cramér’s V measured the strength of these associations.

## Results

### Participants

294 participants were contacted from the KHSC mental health ambulatory clinic general waitlist from January to September 2023. 83 files were closed due to a lack of response (standard hospital policy: three calls, one email) or no longer residing within Ontario, 13 declined cares, and 62 chose not to participate, yielding 136 participants. Of these, five showed interest but did not activate their account, 30 activated their account but did not complete the AI-assisted triage module, and 101 completed the triage (36.05 years (SD=12.17), 71.74% female). Most were Canadian-born (89.13%) and about half had children (45.65%; number of children=2.24 (SD=1.23), age of children=20.38 years (SD=9.84)). Those not born in Canada (n=10) immigrated at 16.50 (SD=15.66) years of age. All participants were fluent in English (Table 1). While 101 participants completed the triage module, only 92 completed the demographic information in the module (Table 1).

**Table 1.** Participant demographics.

| **Demographic Variable** | **n (%)** |
| --- | --- |
| **Sex (n=92)** |  |
| *Female* | 66 (71.74) |
| *Male* | 26 (28.26) |
| **Gender Identity (n=92)** |  |
| *Female* | 61 (66.30) |
| *Male* | 26 (28.26) |
| *Non-Binary* | 3 (3.26) |
| *Other* | 2 (2.17) |
| **Ethnicity (n=92)** |  |
| *White* | 84 (91.30) |
| *Black* | 2 (2.17) |
| *Indigenous* | 2 (2.17) |
| *Hispanic* | 1 (1.09) |
| *Other* | 3 (3.26) |
| **Marital Status (n=92)** |  |
| *Never Married* | 47 (51.09) |
| *Married* | 23 (25.00) |
| *Divorced* | 7 (7.61) |
| *Widowed* | 3 (3.26) |
| *Other* | 12 (13.04) |
| **Highest Level of Education Completed (n=90)** |  |
| *Did not complete secondary school* | 4 (4.44) |
| *Secondary school diploma* | 27 (30.00) |
| *College diploma* | 29 (32.22) |
| *Undergraduate Degree* | 23 (25.56) |
| *Master's Degree* | 5 (5.56) |
| *PhD* | 2 (2.22) |
| **Employment Status (n=92)** |  |
| *Full-Time* | 36 (39.13) |
| *Unemployed* | 23 (25.00) |
| *Part-Time* | 6 (6.52) |
| *Student* | 5 (5.43) |
| *Other* | 22 (23.91) |
| **Annual Income (n=91)** |  |
| *Under \$20,000* | 32 (35.16) |
| *\$20,000-34,999* | 14 (15.38) |
| *\$35,000-49,999* | 17 (18.68) |
| *\$50,000-74,999* | 12 (13.19) |
| *\$75,000-99,999* | 11 (12.09) |
| *Over \$100,000* | 5 (5.49) |

### Wait Time to Receive Care

After completing the triage module, participants were matched with therapists and received their first digital psychotherapy module within 2.64 weeks (SD=2.72). Levels 3 and 4 participants requiring psychiatric assessments were seen within 6.67 weeks (SD=3.18), while Levels 1 and 2 participants needing later psychiatric consultations were seen within 10.90 weeks (SD=3.18). Before this program, the HDH Ambulatory Mental Health and Addiction program had a general stream wait time of seven months and an overall wait time of 14 months. By March 2024, the general stream wait time decreased to two months (71.43% improvement), and the overall wait time dropped to 8.48 months (SD=1.82), a 39.43% improvement.

The AI-assisted triage program optimized patient-clinician interactions. Intake calls averaged 6:23 (SD=4:56) compared to 45 minutes with a nurse in traditional practice. Digital modules reduced therapist time per patient from one hour in face-to-face sessions to 14:42 (SD=4:09) for Level 1 participants, with additional weekly check-ins averaging 12:12 (SD=5:13) for Levels 2 and 3.

### AI-Assisted Triage Program Decision-Making Accuracy

The AI-assisted triage program categorized participants as mild (27/101; 26.73%), moderate (55/101; 54.45%), severe (4/101; 3.96%), and inconclusive (15/101; 14.85%) (Figure 3). It matched psychiatrist classifications for 72/101 (71.29%) participants, including mild (23/29), moderate (43/51), severe (3/18), and inconclusive (3/3) cases. A chi-square test showed significant differences in care intensity classifications (X²(9)=95.9, p<0.0001), with Cramér’s V (V=0.56) indicating a moderate-to-strong association. However, 12 cases classified as severe by psychiatrists were rated as moderate by the program, suggesting conservative severity assessments.

**Figure 3.**
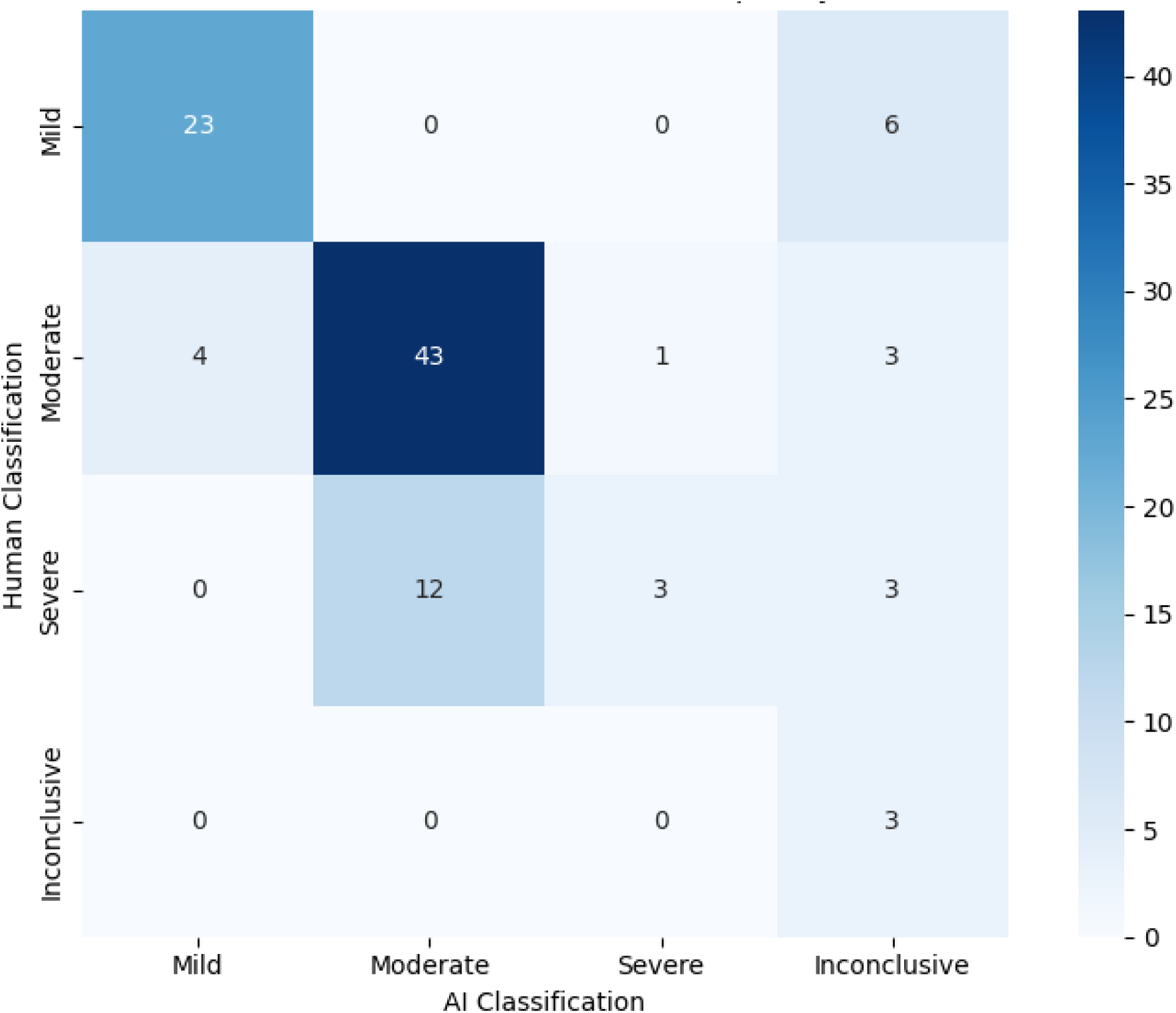
Heat map/crosstab displaying the treatment intensity decision-making of the AI-assisted triage program versus the psychiatrist.

Psychotherapy module recommendations also aligned for 72/101 (71.29%) participants, with a strong agreement for GAD, MDD, DBT, PTSD, COVID-19, and ADHD modules (Figure 4). A chi-square test showed significant classification differences (X²(48)=389, p<0.0001), with Cramér’s V (V=0.8) indicating a moderate-to-strong association. Notably, 7/37 (18.92%) GAD cases were labelled inconclusive by the program, and 0/4 OCD cases were classified correctly, with 3/4 (75.00%) misclassified as DBT. These results suggest challenges in distinguishing GAD and OCD features.

**Figure 4.**
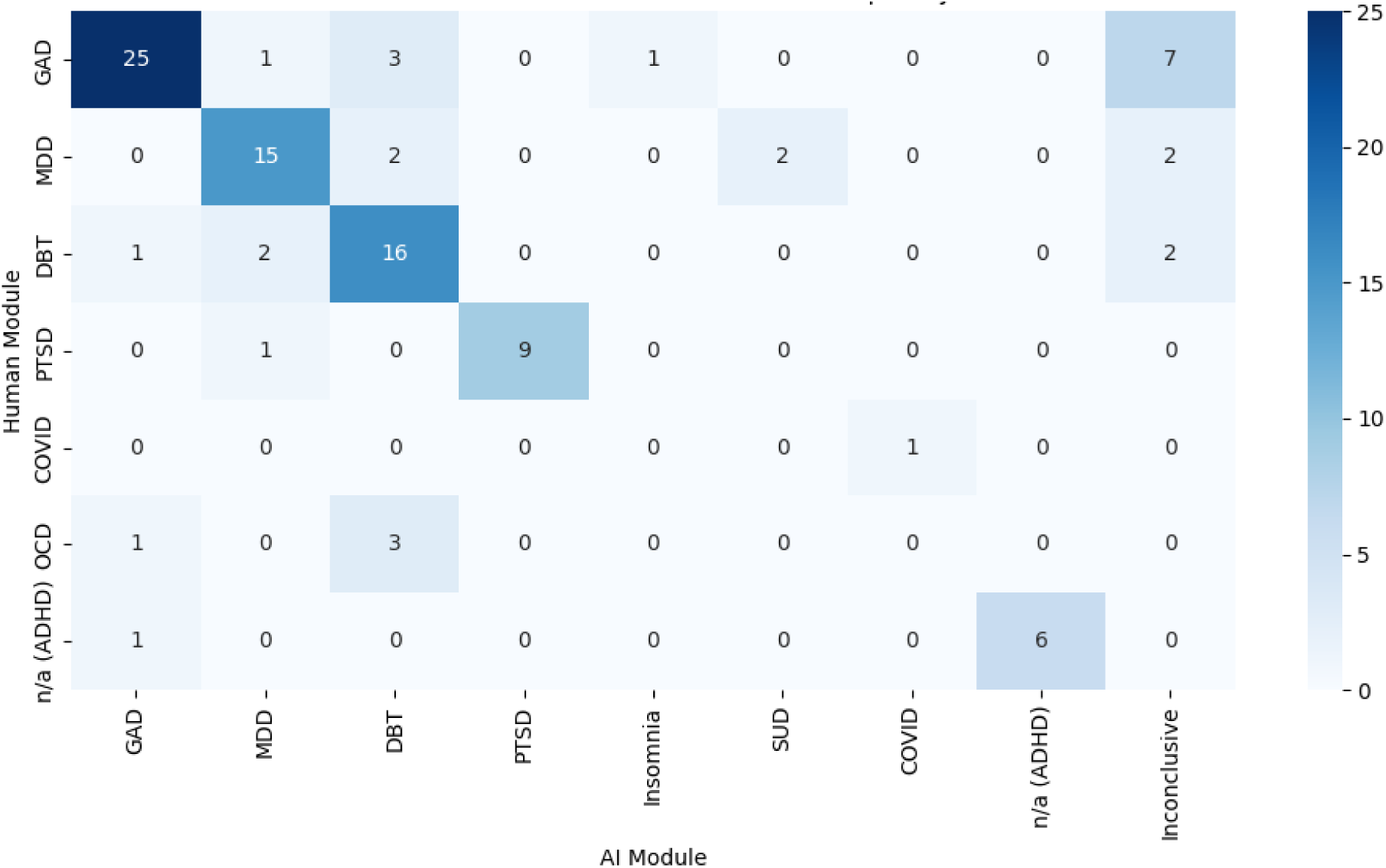
Heat map/crosstab displaying the disorder-specific digital psychotherapy module decision-making of the AI-assisted triage program versus the psychiatrist.

### Psychiatric Consultations

Approximately 24.14% (7/29) and 40.00% (22/50) from Levels 1 and 2 required psychiatric consultation later, resulting in 63.29% of Level 1 and 2 patients not needing to see a psychiatrist. Additionally, all patients flagged for high-risk (n=3) by the clinical team were also flagged by the AI-assisted triage program.

## Discussion

The AI-assisted triage program reduced wait times by streamlining decisions, increasing system capacity with digital psychotherapy, and optimizing resource use. It enabled efficient, coordinated mental healthcare through scalable digital solutions, instantly assessing multiple patients simultaneously. The program prioritized patients requiring psychiatrist access while directing others to online psychotherapy, optimizing resource allocation. Its online format further improved accessibility, scalability, and convenience.

The AI-assisted triage program reduced wait times by over 70%, with patients receiving care within 2.5 weeks of completing the triage module. Severe and inconclusive cases saw a psychiatrist within an average of 6.5 weeks—longer than the 4-week target but still a significant improvement. Mild and moderate cases needing psychiatric care were seen within 11 weeks, meeting the 3–6-month target. Notably, over 60% of mild and moderate cases (half of all participants) did not require psychiatric consultation, optimizing resource allocation. Psychiatrist appointments were reserved for critical cases, while others accessed immediate online psychotherapy, often sufficient for their needs.

The AI-assisted triage program showed strong agreement with psychiatrists (>70%), demonstrating its reliability in recommending care intensity and disorder-specific psychotherapy modules. These findings suggest that AI-assisted triage systems can enhance mental healthcare by improving accessibility and optimizing resources. While using a psychiatrist as a comparator provided a robust benchmark, it’s important to note that triage decisions are inherently subjective. Variability among clinicians would likely occur if multiple experts were involved, highlighting the value of a standardized, evidence-based system^53^. For example, prior research showed a model distinguishing symptomatic from asymptomatic sentences achieved 74% accuracy, comparable to a 76% inter-rater overlap between two human experts^26^. This highlights the potential of AI-assisted triage to reduce inconsistencies and support equitable care delivery.

The AI-assisted triage program exhibited biases in decision-making, underdiagnosing some participants by categorizing those deemed severe by psychiatrists as moderate. These insights will be used to refine the algorithm, underscoring the importance of collaboration between clinicians and AI developers to enhance decision-making tools. While not perfectly accurate, the algorithm ensured patients received some level of care while on the waitlist—preferable to no care at all. Additionally, every patient was assigned a clinician who could reassess and adjust their care level as needed.

This trial did not deeply explore the role of decision confidence in decision-making accuracy. The algorithm suggested multiple relevant digital psychotherapy modules based on patients’ narratives and reported the probability of their relevance (Figure 1). For this study, the module with the highest probability was selected as the AI decision. Preliminary analysis (unreported) indicated that when the top suggestion had a probability >40%, the algorithm’s accuracy exceeded 90%, occurring in 65% of cases. This suggests decision-making accuracy could improve if the algorithm were used as a supportive tool. Clinicians could rely on high-confidence outcomes while directing low-confidence cases for further evaluation.

The real-world clinical setting, where this project was carried out, enhances the generalizability of the findings and demonstrates the feasibility of such systems. However, several limitations should be noted. The lack of long-term follow-up data limits the ability to assess the program’s sustainability and lasting effects on patient outcomes. Furthermore, the findings of this observational study do not have the strength of those that would have been obtained by a randomized design. Selection bias may also have impacted the results, as voluntary participation could have led to a sample not fully representative of the broader patient population. As a novel care delivery model, participants may have faced technical challenges or trust issues with an AI-enabled solution. To address this, a qualitative investigation is underway to explore the challenges, barriers, and perceptions of participants when it comes to the use of AI in mental healthcare and the psychotherapy platform^54^.

Despite these limitations, this research provides a strong foundation for future investigations into AI-assisted triage in mental healthcare. Several areas warrant further exploration. First, assessing the scalability of this program should be investigated. Moreover, the long-term outcomes of these tools, including patient quality of life, functional outcomes, overall well-being, changes in ED utilization, length of hospital stay, rate of readmission, and costs of managing other chronic diseases are crucial to understand. Additionally, replicating these implementations in diverse settings and populations is necessary to examine the generalizability of the findings.

Future research should also include a comprehensive financial analysis to guide healthcare system decisions. Untreated mental health issues impose substantial costs, with healthcare utilization costs being 3.5 times higher for patients with depression, for example, versus those without ($10,064 vs. $2,832) and social service costs being triple ($1,522 vs. $510)^55,56^. Overall, mental health problems can add an average annual cost of $8,244 per person (SD=40,542)^55,56^. Additionally, unemployment represents the highest per capita cost, with an annual employment loss valued at $32,750 per person^55,56^. Through this project, wait times were shortened by five months, saving $17,080/per person ($3,435 in healthcare, $13,645 in productivity). In total, this project (n=101) potentially saved the Canadian economy $1,725,080.

As AI advances, ethical concerns like privacy, data security, and bias must be prioritized to build trust and integrate AI solutions into mental healthcare. Future research should explore these issues to better understand the benefits and limitations of AI-assisted triage, supporting broader implementation and improved patient care.

## Conclusion

This initiative evaluated the effectiveness of an AI-assisted triage program in reducing wait times and delivering personalized, patient-centred care. Diagnosis-specific online psychotherapy programs were offered as accessible, scalable, cost-effective, and validated treatments, enhancing patients’ quality of life. OPTT’s AI-assisted triage program enabled standardized, high-quality care while maximizing system capacity. At a system level, it demonstrated the potential to reduce wait times and improve equitable, affordable care access. By enhancing capacity, streamlining workflows, and supporting clinical decision-making, this model addresses key challenges in care delivery. Additionally, it may reduce ED strain, improve chronic disease management by addressing comorbidities, and establish a foundation for measurement-based care by integrating evidence-based metrics to optimize outcomes.

## Data Access

Data is available as needed upon request.

## Acknowledgements

Thank you to Alina Marin, Archana Patel, Amanda Richer, Behnia Hagiri, Meghanne Hicks, Julia Lee, and Nicholas Axas for their contributions.

## Plain Language Summary

The mental healthcare system is overwhelmed by increasing patient demand, leading to long wait times for care. The needs of each patient vary based on their specific condition and the resources available. Digital health tools and artificial intelligence could help to address these challenges by streamlining care and matching patients to appropriate resources. This study explored the use of an artificial intelligence-assisted triage tool to improve the process of assigning care levels and treatment recommendations for patients waiting for psychiatric care. Researchers compared this artificial intelligence system to the decision-making of a psychiatrist. The study involved 101 patients from an outpatient psychiatry waitlist. Each participant completed a digital triage module, which used natural language processing and machine learning to recommend a care intensity level and suggest a digital psychotherapy program tailored to their needs. The psychiatrist also reviewed the same participants’ responses and compared the results. The findings were promising since they showed that the AI-assisted triage program reduced wait times by 71%, allowing participants to access care within three weeks of completing the module. Additionally, in over 70% of cases, the AI-assisted triage program recommendations for treatment intensity and psychotherapy matched those of the psychiatrist. Furthermore, 63% of patients who were allocated to lower-intensity care plans, by artificial intelligence, did not need further psychiatric consultation. This suggests that the tool effectively identified appropriate care pathways for many patients. These results highlight the potential of artificial intelligence in alleviating the burden on mental healthcare systems. By accelerating triaging and improving decision-making, these digital tools could increase care capacity and streamline workflows. With further refinement, this technology could be a game-changer for hospitals and mental health services.

